# Anti-SARS-CoV-2 antibody levels are concordant across multiple platforms but are not fully predictive of sterilizing immunity

**DOI:** 10.1101/2021.04.26.21256118

**Authors:** Benjamin T. Bradley, Andrew Bryan, Susan L. Fink, Erin A. Goecker, Pavitra Roychoudhury, Meei-Li Huang, Haiying Zhu, Anu Chaudhary, Bhanupriya Madarampalli, Joyce Y.C. Lu, Kathy Strand, Estella Whimbey, Chloe Bryson-Cahn, Adrienne Schippers, Nandita S. Mani, Gregory Pepper, Keith R. Jerome, Chihiro Morishima, Robert W. Coombs, Mark Wener, Seth Cohen, Alexander L. Greninger

## Abstract

With the availability of widespread SARS-CoV-2 vaccination, high-throughput quantitative anti-spike serological testing will likely become increasingly important. Here, we investigated the performance characteristics of the recently FDA authorized semi-quantitative anti-spike AdviseDx SARS-CoV-2 IgG II assay compared to the FDA authorized anti-nucleocapsid Abbott Architect SARS-CoV-2 IgG, Roche elecsys Anti-SARS-CoV-2-S, EuroImmun Anti-SARS-CoV-2 ELISA, and GenScript surrogate virus neutralization assays and examined the humoral response associated with vaccination, natural protection, and breakthrough infection. The AdviseDx assay had a clinical sensitivity at 14 days post-symptom onset or 10 days post PCR detection of 95.6% (65/68, 95% CI: 87.8-98.8%) with two discrepant individuals seroconverting shortly thereafter. The AdviseDx assay demonstrated 100% positive percent agreement with the four other assays examined using the same symptom onset or PCR detection cutoffs. Using a recently available WHO International Standard for anti-SARS-CoV-2 antibody, we provide assay unit conversion factors to international units for each of the assays examined. We performed a longitudinal survey of healthy vaccinated individuals, finding median AdviseDx immunoglobulin levels peaked seven weeks post-first vaccine dose at approximately 4,000 IU/mL. Intriguingly, among the five assays examined, there was no significant difference in antigen binding level or neutralizing activity between two seropositive patients protected against SARS-CoV-2 infection in a previously described fishing vessel outbreak and five healthcare workers who experienced vaccine breakthrough of SARS-CoV-2 infection – all with variants of concern. These findings suggest that protection against SARS-CoV-2 infection cannot currently be predicted exclusively using *in vitro* antibody assays against wildtype SARS-CoV-2 spike. Further work is required to establish protective correlates of protection for SARS-CoV-2 infection.

## Introduction

Severe acute respiratory syndrome coronavirus 2 (SARS-CoV-2) the etiologic agent of coronavirus disease 19 (COVID-19) is responsible for an ongoing global pandemic. In addition to infection control measures such as social distancing and masking, controlling the spread of the outbreak will require a global vaccination campaign. Currently, three vaccines have received FDA emergency use authorization with other candidates in late-phase clinical trials (1). Common to all vaccine candidates is the inclusion of the receptor binding domain (RBD) or full-length spike (S) of SARS-CoV-2 (2).

For SARS-CoV-2 and related coronaviruses, antibodies to the RBD of the spike protein have demonstrated potent neutralizing activity at nanomolar concentrations (3, 4). In a recent meta-analysis of individuals with naturally-acquired SARS-CoV-2 infection, neutralizing antibodies were first detectable between seven to 15 days following symptom onset (5). Despite questions regarding the durability of the antibody response and documented cases of reinfection, longitudinal analysis of IgG levels and neutralizing potency suggest immunity persists in most individuals for as long a time period as has been examinable to date (6, 7). However, some individuals, including those who are older or immunosuppressed, may be at risk for a suboptimal response to vaccination (8, 9).

The presence of neutralizing antibodies due to prior infection or vaccination has been shown to be a correlate of protection against SARS-CoV-2 infection (10–12). Although Phase III vaccine trials demonstrated excellent efficacies among treated populations (13, 14), a number of subpopulations including pregnant women and immunocompromised individuals were excluded from these trials. Moreover, uncertainty exists over the durability of protection after vaccination (15). High-throughput, widely available laboratory measurements of protective correlates would be extremely helpful in these and other populations. The current gold-standard test, known as the plaque reduction neutralization assay (PRNA), is resource intensive and requires BSL-3 conditions for testing. Currently, one surrogate neutralization assay has received EUA-approval for clinical use. While this assay can be performed in BSL-2 laboratories and has shown excellent correlation to PRNAs, it also suffers from similar limitations of throughput and cost (16). The most widely used clinical platforms for monitoring immunity to vaccine-preventable diseases including hepatitis B virus, measles virus, and varicella-zoster virus are high-throughput, low-cost immunoassay analyzers, including the Roche cobas, Abbott Architect, and Diasorin XL platforms, among others. Recently, the Abbott AdviseDx SARS-CoV-2 IgG II assay received emergency use authorization by the FDA. This chemiluminescent microparticle immunoassay (CIMA) for the Abbott Architect platform is designed for semi-quantitative detection of IgG class antibodies to the RBD of the SARS-CoV-2 spike protein.

Our laboratory previously examined the clinical performance characteristics of the anti-N SARS-CoV-2 IgG assay for the Abbott Architect and found it to have adequate performance for determining prior SARS-CoV-2 infection in a hospitalized cohort (17). However, this assay is qualitative and designed to detect antibodies to the nucleocapsid, precluding the ability to monitor vaccine response. In this study we examine the performance of the AdviseDx SARS-CoV-2 IgG II assay and correlate its performance to four other assays (Abbott Architect SARS-CoV-2 anti-nucleocapsid IgG, Roche Elecsys Anti-SARS-CoV-2 S, EuroImmun Anti-SARS-CoV-2 ELISA IgG, and GenScript surrogate virus neutralization test). The WHO International Standard was also run on each platform to evaluate analytical sensitivity using the manufacturer’s cutoffs.

Important questions remain as to what binding antibody levels may be considered protective against SARS-CoV-2 infection. We explored this question using the following approach: First, we calculated median immunoglobulin values following SARS-CoV-2 vaccination over 11 weeks in a group of healthy volunteers. Second, we compared the antibody response between a pair of seropositive patients who were protected from SARS-CoV-2 infection during a previously described outbreak on a fishery vessel and a group of five fully vaccinated healthy individuals who subsequently experienced breakthrough SARS-CoV-2 infection. This study not only demonstrates the acceptable analytical clinical performance of the Abbott AdviseDx assay, but also provides context for how these values may be interpreted in vaccinated individuals.

## Materials and Methods

### Study population & Specimen collection

A total of 128 residual plasma specimens from 91 patients with a history of PCR-confirmed SARS-CoV-2 infection or anti-N IgG antibodies were included in this study (raw data in Supplemental Dataset 1). Residual serum from 104 individuals collected between June and August of 2019 for anti-HSV western blot analysis was used as the negative control. 155 samples from 27 vaccinated asymptomatic ambulatory adult health care workers were collected to assess the longitudinal response (raw data available in Supplemental Dataset 2. These individuals received either the mRNA-1273 or BNT162b2 vaccine. Samples were obtained from two seropositive individuals who previously demonstrated protection against SARS-CoV-2 infection during a previously described fishery vessel outbreak (10). Five samples were obtained from fully vaccinated healthcare workers with PCR-confirmed SARS-CoV-2 infection and mild COVID-19 symptoms. Diagnostic SARS-CoV-2 PCR tests included a Washington state authorized CDC-based laboratory developed assay or FDA authorized Roche cobas SARS-CoV-2, Abbott Alinity m SARS-CoV-2, or Hologic Panther Fusion SARS-CoV-2 assays (18). SARS-CoV-2 Whole genome sequencing was performed using the Swift Biosciences v2 or Illumina COVID-Seq amplicon tiling assays (19). This study was approved by the University of Washington Institutional Review Board.

### Anti-N Abbott Architect SARS-CoV-2 IgG and Anti-S AdviseDx SARS-CoV-2 IgG II assays

The emergency use authorized anti-N Architect SARS-CoV-2 IgG and anti-S AdviseDx SARS-CoV-2 IgG II assays (Abbott, Chicago, IL) are chemiluminescent microparticle immunoassay (CIMA) designed to measure IgG antibodies binding the N protein and S protein, respectively, and were performed on an Architect i2000SR. Results from the anti-N SARS-CoV-2 IgG assay are reported as index values. The index value of 1.40 or greater was classified as positive per manufacturer’s recommendation for the anti-N Abbott Abbott Architect SARS-CoV-2 IgG assay. Results from the anti-S AdviseDx SARS-CoV-2 IgG II assay are reported as arbitrary units per milliliter (AU/mL). The manufacturer’s suggested positive cutoff of 50 AU/mL was used. Inter-day and inter-assay studies were performed over three days by two different operators.

### Roche Elecsys Anti-SARS-CoV-2 S

The Elecsys Anti-SARS-CoV-2 S assay (Roche Diagnostics International Ltd, Rotkreuz, Switzerland) is an electrochemiluminescence immunoassay which uses a double-antigen sandwich design for the detection of immunoglobulins (predominantly IgG, but also IgA and IgM) to the RBD domain of the S protein. Samples were prepared according to the manufacturer’s instructions and analyzed on the Roche cobas e 411 platform. Using the manufacturer’s guidelines, sample values ≥0.8 AU/mL were classified as positive for anti-SARS-CoV-2 antibodies. No dilutions were performed on specimens to extend the measurement range of the assay which does not report values above 250 AU/mL.

### EuroImmun Anti-SARS-CoV-2 ELISA

The EuroImmun anti-SARS-CoV-2 IgG assay is a semi-quantitative ELISA detecting antibodies which bind the S1 subunit of the spike protein. Samples are loaded into reagent wells coated with the spike protein, washed, and then incubated with enzyme-conjugated anti-human IgG generating a colorimetric signal. Results are provided as a semi-quantitative measurement of the signal of the experimental sample divided by the signal of the calibrator (OD ratio). Per the manufacturer’s insert values < 0.8 are considered negative, ≥ 0.8 to <1.0 borderline, and ≥ 1.1 positive. For this study we classified borderline results as positive.

### GenScript Surrogate Virus Neutralization Test

The GenScript Surrogate Virus Neutralization Test (Piscataway, NJ, USA) assay was performed according to the manufacturer’s instructions. The test examines the ability of sera to block binding of SARS-CoV-2 spike RBD to the human ACE2 receptor. Absorbance was read at 450 nm on a VICTOR Nivo (PerkinElmer, Waltham, MA, USA) reader. Values are reported as percent neutralization relative to a negative control sample provided by the manufacturer. Samples demonstrating ≥30% inhibition of ACE2 binding were classified as positive as recommended by the manufacturer.

### SARS-CoV-2 Spike Pseudotyped Lentivirus Neutralization assay

1.25E^4^ 293T-ACE2 cells were seeded in 96-well plates and incubated in DMEM + 10% FBS for 16-18 hrs. The following day, 1.0E^7^ RLUs/well SARS-CoV-2 D614G Spike pseudotyped lentivirus was diluted 1:10 in DMEM with 10% FBS complete media (20). Sera was diluted 1:20 in DMEM with 10% FBS and seven three-fold serial dilutions were prepared. Equal parts diluted sera and pseudovirus were combined and incubated for one hour at 37°C. The mixture was added to the cells and incubated for fifty-two hours at 37°C. Following incubation, the media was removed and 30µl of luciferase substrate (Promega, Madison, WI, USA) was added. After two minutes of incubation, luminescence was measured on the VICTOR Nivo and IC50 was calculated from a standard curve using the CV30 monoclonal antibody (Absolute Antibody, Oxford, UK).

### Preparation of the International Standard

The WHO First International Standard for SARS-CoV-2 antibody were prepared according to the manufacturer’s instructions (21). The lyophilized sample was provided at 250IU/ampoule and resuspended in 250µl of deionized water to create a 1000 IU/mL stock solution. The stock was diluted 1:10 to prepare a working solution with sufficient volume for analysis across the various platforms.

### Statistical Analysis

Correlation studies were performed using Spearman’s coefficient. Assay performance, linear regression, curve fitting calculations were performed using Prism 9 (GraphPad Software, LLC, San Diego, CA, USA).

## Results

### The AdviseDx SARS-CoV-2 IgG II assay is 95.6% sensitive and 100% specific in individuals at least 14 days post-symptom onset or 10 days post-first positive PCR result

To assess the sensitivity and specificity of the AdviseDx SARS-CoV-2 IgG II assay, a total of 172 patient samples (68 positive, 104 negative) were tested. Positive cases were classified as individuals with a PCR-confirmed diagnosis of SARS-CoV-2 infection who were at least 14 days post-symptom onset or 10 days post-first positive PCR result. Negative sera were collected during July and August 2019, prior to when SARS-CoV-2 was thought to be circulating in the western Washington area (22). Using these criteria, the assay had a sensitivity of 95.6% (65/68 samples; 95% CI: 87.8-98.8%) (Figure 1A, Supplemental Table 1). Importantly, two of the three cases that initially tested negative were in clinically asymptomatic individuals detected by pre-admission SARS-CoV-2 PCR screening. Based on the manufacturer’s recommended cutoff of 50 AU/mL, these two individuals tested antibody negative on days 11 (13.8 AU/mL) and 13 (10.9 AU/mL) and seroconverted on days 13 (77.3 AU/mL) and 18 (168.3 AU/mL) post-PCR, respectively. The third individual was borderline anti-S seronegative (19.5 AU/mL) at day 15 post-symptom onset and died due to COVID-19 pneumonia on day 17 post-symptom onset. Specificity was calculated to be 100% (104/104 samples; 95% CI 96.4-100%) (Table 1B). The median AU/mL of these specimens was 1.8 AU/mL (range 0 – 32.9 AU/mL), and the limit of blank, calculated as the mean of negative samples + 3 S.D., was 17.3 AU/mL, well below the manufacturer’s recommended cutoff of 50 AU/mL.

**Table 1.**
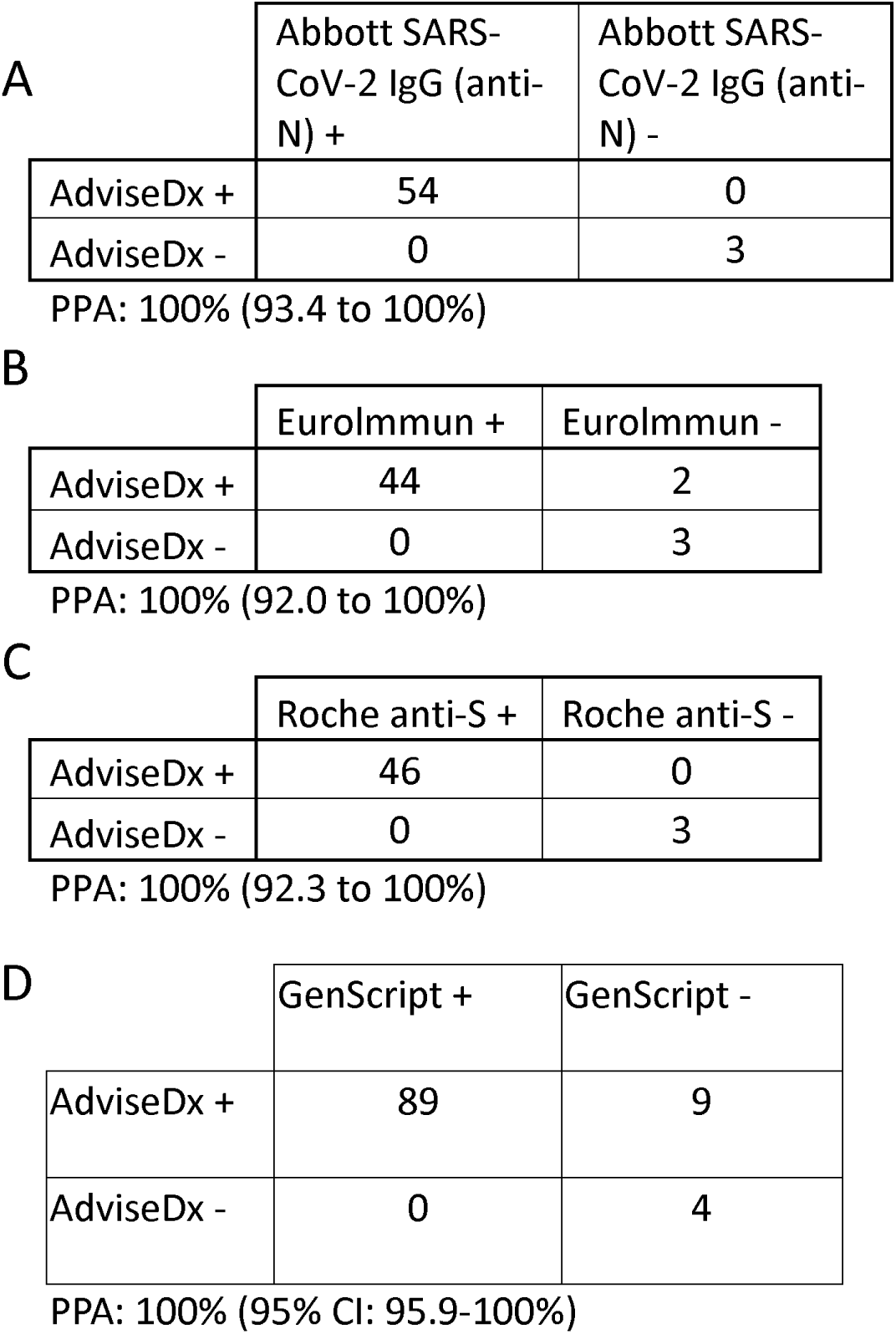
Excellent categorical agreement is observed between the AdviseDx SARS-CoV-2 IgG II assays and Abbott SARS-CoV-2 IgG (A), EuroImmun (B), and Roche Elecsys Anti-SARS-CoV-2 S (C) enzyme immunoassays. Positive percent agreement was calculated for samples from patients that were greater than 14 days from symptom onset or 10 days post first positive PCR result. Positive percent agreement of the AdviseDx SARS-CoV-2 IgG II assays to the GenScript surrogate virus neutralization was 100% when all samples with GenScript data were measured irrespective of collection time (D). In discrepant cases (AdviseDx+/ GenScript-) the AdviseDx result ranged from 50.1-290.7 AU/mL, above the assay positive threshold of 50 AU/mL.

|  | Abbott SARS-CoV-2 IgG (anti-N) + | Abbott SARS-CoV-2 IgG (anti-N) - |
| --- | --- | --- |
| AdviseDx + | 54 | 0 |
| AdviseDx - | 0 | 3 |

PPA: 100% (93.4 to 100%)

B
|  | Eurolmmun + | Eurolmmun - |
| --- | --- | --- |
| AdviseDx + | 44 | 2 |
| AdviseDx - | 0 | 3 |

PPA: 100% (92.0 to 100%)

C
|  | Roche anti-S + | Roche anti-S - |
| --- | --- | --- |
| AdviseDx + | 46 | 0 |
| AdviseDx - | 0 | 3 |

PPA: 100% (92.3 to 100%)

**Table 1** Excellent categorical agreement is observed between the AdviseDx SARS-CoV-2 IgG II assays and Abbott SARS-CoV-2 IgG (A), EuroImmune (B), and Roche Elecsys Anti-SARS-CoV-2 S (C) enzyme immunoassays.
|  | GenScript + | GenScript - |
| --- | --- | --- |
| AdviseDx + | 89 | 9 |
| AdviseDx - | 0 | 4 |

PPA: 100% (95% CI: 95.9-100%)

**Figure 1.**
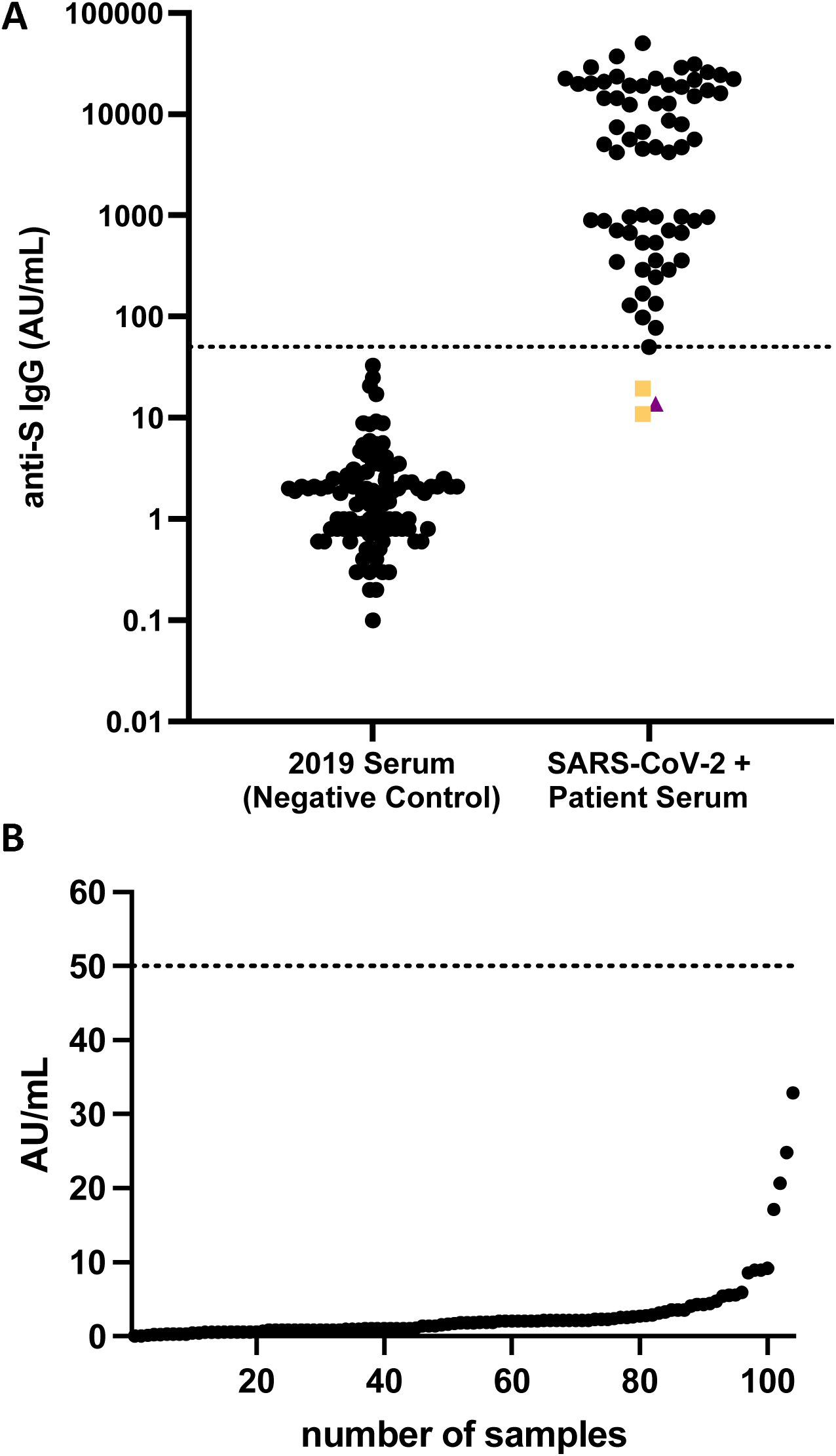
AdviseDx SARS-CoV-2 IgG II values for patients with a PCR-confirmed SARS-CoV-2 infection who were >14 days post symptom onset or >10 days following first positive PCR result and control sera collection prior to the SARS-CoV-2 pandemic (A). The dotted line represents the assay positive cutoff (50 AU/mL) set by the manufacturer. Gold squares indicate patients who seroconverted at 13 and 18 days following their first positive PCR result. Purple triangle represents a severely ill COVID-19 patient who died 17 days post-symptom onset. 104 serum samples obtained prior to the emergence of SARS-CoV-2 were analyzed for assay specificity (B). The median value of these negative controls was 1.8 AU/mL (range: 0-32.9). The dotted line represents the positive threshold.

### The AdviseDx SARS-CoV-2 IgG II assay is linear over the analytic measurement interval with a coefficient of variation <5% near the positive cutoff

Per the manufacturer’s insert the AdviseDx SARS-CoV-2 IgG II has a limit of quantitation of 22.0 AU/mL and an upper limit of quantitation of 25,000 AU/mL. To assess the linearity of the assay, we performed 1:2 serial dilutions of a high-positive sample from 37,256 AU/mL to ∼12 AU/mL. Each sample dilution was measured in triplicate. The AdviseDx level of the neat sera was calculated from a 10x dilution. Results demonstrated excellent linearity beyond the manufacturer’s analytical measurement interval (R^2^=0.9989) (Supplemental Figure 1). To assess for assay reproducibility, the coefficient of variation (CV) was measured in four samples near the manufacturer’s suggested positive cutoff over four days. The assay demonstrated a CV less than 5% at all dilutions above the positive threshold (550, 140, and 70 AU/mL) during intraday and interday measurements. For the sample below the positive threshold (40 AU/mL) the CV was 5% and 7.7% on intraday and interday measurements, respectively (Supplemental Table 2).

### The AdviseDx SARS-CoV-2 IgG II has 100% positive agreement with three other EUA immunoassays

Results of the AdviseDx assay were compared to three serologic binding assays with prior EUAs, Abbott Architect SARS-CoV-2 IgG (nucleocapsid), EuroImmun Anti-SARS-CoV-2 ELISA (spike S1 subunit), and Roche Elecsys Anti-SARS-CoV-2 S (spike RBD). Using a cutoff of 14 days post-symptom onset or 10 days post first positive PCR result, the positive percent agreement (PPA) of the AdviseDx with the three other assays was 100% (Table 1 A-C). When patient samples were examined regardless of collection time, the AdviseDx PPA agreement for the Abbott Architect, EuroImmun, and Roche assays were 98.3% (114/116), 100% (95/95), and 100% (102/102) respectively (Supplemental Table 3).

A total of 13 samples with at least one qualitative test result discrepancy were observed, which were mostly driven by negative results on the EuroImmun test (Supplemental Table 4). Discrepant cases occurred near the time of seroconversion in patients with available clinical data. Of the discrepant cases, two were considered to be early positives by the Abbott Architect anti-N assay based on negative results on the other three platforms. Both of these cases had Abbott Architect anti-N index values close the cutoff at 1.53 and 1.44. Of the remaining cases, 11/11 were AdviseDx positive, 10/11 Abbott Architect anti-N positive, 8/11 Roche positive, and 0/11 EuroImmun positive. Quantitative results of the AdviseDx assay were highly correlated to the three other platforms as measured by Spearman’s coefficient: Abbott Architect anti-N (r=0.89), EuroImmun anti-S1 (r=0.95), Roche anti-RBD (r=0.90), p<0.001 for all comparisons. Linear regression and goodness of fit were calculated following log-transformation of AdviseDx values. The goodness of fit (R^2^) of the log-transformed AdviseDx values to the quantitative results of the Abbott Architect anti-N, EuroImmun anti-S1, and Roche anti-S/RBD assays were 0.42, 0.84, and 0.73, respectively (Figure 2 A-C), which was chiefly affected by the limited reportable range of these assays. Categorical agreement and assessment of linearity between the Abbott Architect anti-N, EuroImmun, Roche, and GenScript assays are available in Supplemental Figure 2 and Supplemental Table 5.

**Figure 2.**
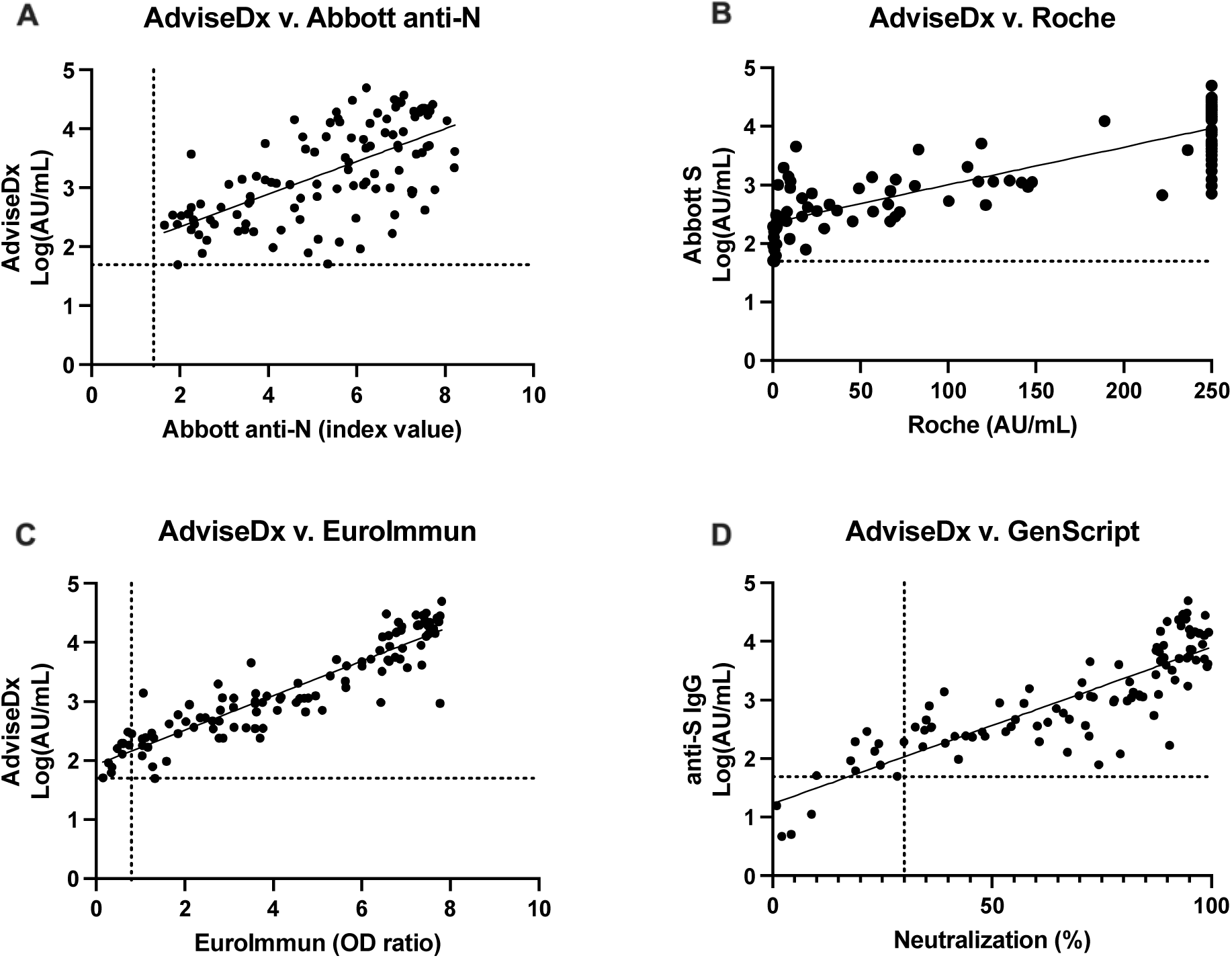
Correlation of AdviseDx SARS-CoV-2 IgG II assays results to the Abbott Architect anti-N (A), EuroImmun anti-S (B), Roche anti-S (C), and GenScript (D) assays. The same specimens were run on all five platforms. AdviseDx results were log-transformed before performing linear regression. Results demonstrated strong agreement and goodness of fit (R^2^) was measured as 0.42, 0.84, 0.73, and 0.74, respectively. Dotted lines represent the manufacturer’s positive cutoff value for each assay.

### The AdviseDx SARS-CoV-2 IgG II assay demonstrates 100% positive agreement with FDA-emergency use authorized surrogate virus neutralization test

As the GenScript Surrogate Virus Neutralization Test is the only currently available neutralization-based assay that has been authorized by the FDA, we compared its results to the AdviseDx SARS-CoV-2 IgG II results. Categorical evaluation demonstrated a positive precent agreement of 100% (89/89 samples; 95% CI: 95.9-100%) in patients with a documented history of SARS-CoV-2 infection (Table 1D). AdviseDx values positively correlated with increasing percent neutralization by Spearman’s analysis (r=0.86, p<0.001). Following log-transformation of the AdviseDx values, a linear regression model was fit to the data (Figure 2D). Neutralization values of 30% (positive cutoff), 50%, and 80% on the GenScript assay corresponded to 107, 369, and 2340 AU/mL on the Abbott IgG II assay, respectively. Similarly, a 18% neutralization value correlated to 50 AU/mL, the positive cutoff, for the AdviseDx assay.

### Increases in anti-S binding antibodies and neutralization activity are observed in individuals following SARS-CoV-2 infection and vaccination

The kinetics of the Genscript and AdviseDx assays were measured over 10-15 days in four patients hospitalized for COVID-19 and over 44-59 days in four individuals who received mRNA vaccines to SARS-CoV-2. Both anti-S immunoglobulins and neutralization values increased over time with high neutralizing levels achieved in all patients (Figure 3). Patients 1 and 2 had positive AdviseDx results on days 8-9, with a positive GenScript 1-2 days later. Patient 4 tested positive by both assays ten days post-symptom onset. For Patient 3, clinical samples were available starting on day 5 post-symptom onset, and both were found to be positive at that time. In the vaccinated patients, a decrease in AdviseDx values was observed at later timepoints although the surrogate neutralizing results remained elevated.

**Figure 3.**
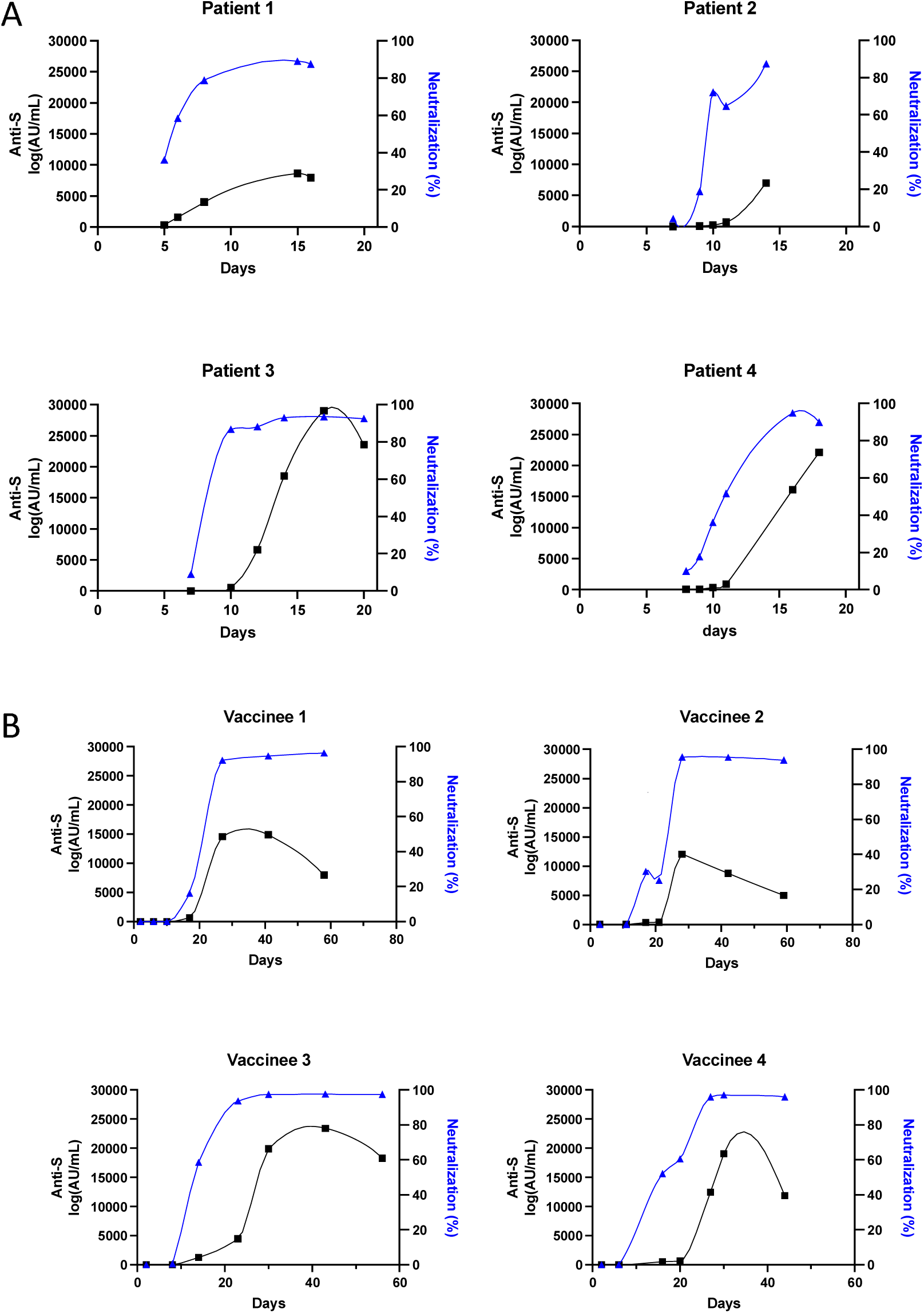
Serial measurements of anti-S IgG levels and surrogate neutralization results in four patients with acute SARS-CoV-2 infection (A) and four patients who received the mRNA-1273 or BNT162b2 SARS-CoV-2 vaccine (B). Increasing anti-S levels and neutralization were observed in all patients as time from exposure increased. At later time points, vaccinated patients had lower anti-S levels with sustained neutralization activity. Curves plotted in Prism 9 using akima spline.

### Implementation of the International Standard demonstrates variable sensitivity among serologic assays for SARS-CoV-2 using the manufacturer’s recommended cutoffs

To determine the positivity threshold for each assay in standardized International Units, a dilution series of the WHO International Standard for SARS-CoV-2 Antibody was prepared and run in triplicate on all platforms (Table 2). For those assays with demonstrated acceptable linearity across the analytic measurement range (AdviseDx, EuroImmun, Roche, Abbott Architect anti-N) the positive cutoff in IU/mL was calculated by linear regression. In increasing order, the manufacturers’ recommended assay cutoffs were determined to be: Roche (3.2 IU/mL), AdviseDx (8.4 IU/mL), EuroImmun (30.2 IU/mL), GenScript (10-50 IU/mL), and Abbott Architect anti-N (56.2 IU/mL). All assays demonstrated an acceptable coefficient of variation (less than 20%) at values near their positive cutoffs.

**Table 2.** Comparison of WHO International Standard results across multiple platforms demonstrates varying levels of sensitivity. The positive cutoff for the Abbott Architect anti-N, AdviseDx, Roche, and EuroImmun assays were calculated as 56.2 IU/mL, 8.4 IU/mL, 3.2 IU/mL, and 30.2 IU/mL, respectively. Each sample was analyzed in triplicate. Positive cutoff: Abbott Architect anti-N (1.4), AdviseDx (50 AU/mL), GenScript (30%), Roche (0.8 AU/mL), EuroImmun (0.8).

| IU/mL | Abbott Architect anti-N (index) |  | AdviseDx anti-S (AU/mL) |  | GenScript (% neutralization) |  | Roche (AU/mL) |  | EuroImmun (OD ratio) |  |
| --- | --- | --- | --- | --- | --- | --- | --- | --- | --- | --- |
|  | mean (%CV) | categorical | mean (%CV) | categorical | mean (%CV) | categorical | mean (%CV) | categorical | mean (%CV) | categorical |
| 100 | 2.5 (1.2%) | POS | 618.9 (0.9%) | POS | 48.3 (3.4%) | POS | 88.2 (1.0%) | POS | 2.30 (2.5%) | POS |
| 50 | 1.27 (1.2%) | NEG | 301.6 (0.6%) | POS | 36.6 (13.6%) | POS | 40.3 (0.1%) | POS | 1.29 (3.1%) | POS |
| 10 | 0.2 (0%) | NEG | 59.5 (4.6%) | POS | 3.9 (78.5%) | NEG | 6.2 (0.1%) | POS | 0.46 (14.6%) | NEG |
| 5 | 0.09 (0%) | NEG | 29.9 (1.0%) | NEG | 3.5 (108%) | NEG | 2.4 (0.03%) | POS | 0.2 (22.9%) | NEG |
| 1 | 0.01 (43%) | NEG | 6.1 (5.7) | NEG | 0.4 (1032.8%) | NEG | 0.4 (0%) | NEG | 0.08 (12.5%) | NEG |

### Longitudinal measurement of anti-SARS-CoV-2 immunoglobulins following vaccination

To understand how anti-S levels change in healthy individuals during vaccination, we examined a total of 155 weekly samples obtained from 27 volunteers (age range 20-72) over up to 11 weeks post-vaccination with either the mRNA-1273 or BNT162b2 SARS-CoV-2 vaccines. Sera were analyzed by the AdviseDx, EuroImmun, and Roche anti-S assays and median values along with the 10^th^-90^th^ percentiles were calculated for the assay results (Table 3). Sera were universally positive by all assays by week 3 after the first vaccine dose. Median AdviseDx anti-spike IgG levels increased more than 100-fold more than two weeks after the first dose and more than 10-fold after administration of the second dose and peaked at seven weeks after first vaccine dose at 23,881 AU/mL (7,304 to >25,000 AU/mL, 10^th^-90^th^ percentiles). All sera obtained more than six weeks after vaccination exceeded the analytical reportable range of the anti-S Roche assay at >250 AU/mL.

**Table 3.** Longitudinal antibody response in a group of healthy volunteers following vaccination with mRNA-1273 or BNT162b2 SARS-CoV-2 as measured by three different anti-S assays. Initial detectable positive values were identified at Week 3 with values peaking at Week 7, consistent with anamnestic booster response. Values are reported as the median antibody levels along with the 10-90^th^ percentile

|  | Abbott AdviseDx (anti-S) |  |  | Roche Elecsys (anti-S) |  |  | Euroimmun (anti-S) |  |  |
| --- | --- | --- | --- | --- | --- | --- | --- | --- | --- |
|  | Median (AU/mL) | 10th Percentile | 90th Percentile | Median (AU/mL) | 10th Percentile | 90th Percentile | Median (index) | 10th Percentile | 90th Percentile |
| Week 1 (n=35) | 2.2 | 0.7 | 4.7 | 0.4 | 0.4 | 0.4 | 0.2 | 0.1 | 0.3 |
| Week 2 (n=22) | 5.2 | 1.4 | 448.3 | 1.8 | 0.4 | 147.8 | 0.3 | 0.2 | 4.2 |
| Week 3 (n=24) | 794.3 | 303.2 | 3823.9 | 40.8 | 8.9 | 227.0 | 4.4 | 2.8 | 9.1 |
| Week 4 (n=22) | 2109.9 | 508.8 | 23958.1 | >250 | 18.1 | >250 | 7.0 | 4.4 | 9.7 |
| Week 5 (n=16) | 21962.1 | 6283.7 | >25000 | >250 | 75.8 | >250 | 9.5 | 5.8 | 10.5 |
| Week 6 (n=13) | 16919.1 | 5200.7 | >25000 | >250 | >250 | >250 | 10.5 | 9.3 | 11.7 |
| Week 7 (n=8) | 23881.4 | 7304.4 | >25000 | >250 | >250 | >250 | 10.6 | 10.0 | 11.4 |
| Week 8 (n=3) | 14648.2 | 7439.6 | 22929.6 | >250 | >250 | >250 | 9.8 | 8.9 | 10.4 |
| Week 9 (n=8) | 8747.2 | 4049.9 | >25000 | >250 | >250 | >250 | 9.4 | 7.8 | 11.4 |
| Week 10-11 (n=4) | 21770.3 | 15256.8 | >25000 | >250 | >250 | >250 | 11.6 | 11.2 | 12.4 |

### SARS-CoV-2 anti-S immunoglobulin levels and neutralization results do not significantly differ between individuals protected after exposure and vaccine breakthrough cases

Prior to the widespread emergence of SARS-CoV-2 variants, we previously described an outbreak of SARS-CoV-2 aboard a fishing vessel in which three individuals with pre-existing SARS-CoV-2 antibodies demonstrated immunity to re-infection (10). These individuals most closely represent the results of a human infection challenge model where an individual with known immunity experiences multiple known exposures to a pathogen. Residual samples were available for two of these individuals with AdviseDx values of 5,303 AU/mL and 1,240 AU/mL. These samples were also tested using across the other available platforms in our study; however, the sample for Patient 2 was exhausted during this process. For the three assays which demonstrated linearity across the analytical range (AdviseDx, EuroImmun, and Roche), the international units per milliliter (IU/mL) were calculated by linear regression. Antibody levels and neutralizing activity were well above the positive cutoff as measured on all platforms, including anti-N (Abbott Architect) and anti-S (AdviseDx, EuroImmun, Roche) antibody responses. When results were standardized to international units, for the three assays which demonstrated linearity across the analytical range (AdviseDx, EuroImmun, and Roche), protection could be observed at levels as low as 81 IU/mL (Table 4).

**Table 4.**
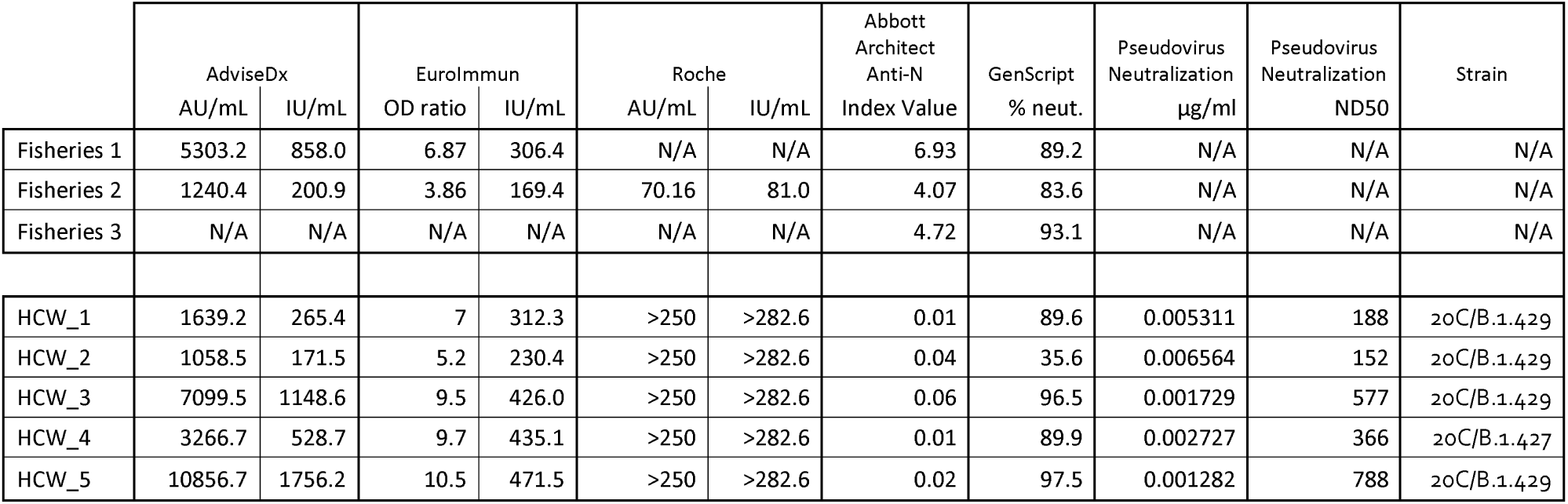
Comparison of SARS-CoV-2 IgG results among three individuals with demonstrated resistance to multiple SARS-CoV-2 challenges and five individuals >6 weeks following vaccination who subsequently developed SARS-CoV-2 infection. Both individuals with resistance to multiple infectious challenge (Fisheries) and vaccine-breakthrough individuals (HCW) demonstrated similar levels of anti-S titers and activity. In only one of five vaccine-breakthrough cases were AdviseDx levels and GenScript neutralization values lower than fishing vessel individuals. International units per milliliter (IU/mL) were calculated for the AdviseDx, Euroimmune, and Roche assays. As expected, Abbott anti-nucleocapsid IgG levels were negative among vaccinated health care workers because they were tested prior to seroconversion to their breakthrough infection, indicating the anti-S levels measured are due to vaccination.

|  | AdviseDx |  | EuroImmun |  | Roche |  | Abbott<br>Architect<br>Anti-N | GenScript | Pseudovirus<br>Neutralization | Pseudovirus<br>Neutralization | Strain |
| --- | --- | --- | --- | --- | --- | --- | --- | --- | --- | --- | --- |
|  | AU/mL | IU/mL | OD ratio | IU/mL | AU/mL | IU/mL | Index Value | % neut. | µg/ml | ND50 |  |
| Fisheries 1 | 5303.2 | 858.0 | 6.87 | 306.4 | N/A | N/A | 6.93 | 89.2 | N/A | N/A | N/A |
| Fisheries 2 | 1240.4 | 200.9 | 3.86 | 169.4 | 70.16 | 81.0 | 4.07 | 83.6 | N/A | N/A | N/A |
| Fisheries 3 | N/A | N/A | N/A | N/A | N/A | N/A | 4.72 | 93.1 | N/A | N/A | N/A |
| HCW_1 | 1639.2 | 265.4 | 7 | 312.3 | >250 | >282.6 | 0.01 | 89.6 | 0.005311 | 188 | 20C/B.1.429 |
| HCW_2 | 1058.5 | 171.5 | 5.2 | 230.4 | >250 | >282.6 | 0.04 | 35.6 | 0.006564 | 152 | 20C/B.1.429 |
| HCW_3 | 7099.5 | 1148.6 | 9.5 | 426.0 | >250 | >282.6 | 0.06 | 96.5 | 0.001729 | 577 | 20C/B.1.429 |
| HCW_4 | 3266.7 | 528.7 | 9.7 | 435.1 | >250 | >282.6 | 0.01 | 89.9 | 0.002727 | 366 | 20C/B.1.427 |
| HCW_5 | 10856.7 | 1756.2 | 10.5 | 471.5 | >250 | >282.6 | 0.02 | 97.5 | 0.001282 | 788 | 20C/B.1.429 |

A second group of samples were collected from five healthcare workers who experienced mild SARS-CoV-2 infection after more than four weeks following their second vaccine dose. All individuals endorsed some form of upper respiratory symptoms after receiving the BNT162b2 vaccine and had surprisingly strong viral loads with an average Ct of 18.4 (range 16.0-20.8, Supplemental Table 6). Serum specimens were collected 1-4 days following symptom onset, tested across all platforms, and standardized to IU/mL when applicable (Table 4). Consistent with a history of prior vaccination and early infection, detectable anti-S and undetectable anti-N antibody responses were observed. In four of five patients, antibody levels and surrogate neutralization results were equal to or greater than samples from the fishing vessel cohort. One patient (HCW_02) demonstrated low positive surrogate neutralization (35.6%, positive cutoff: 30%) and low AdviseDx values compared to the other members of the cohort. However, this patient’s anti-S antibody binding activity as measured on the EuroImmun and Roche platforms were greater than at least one of the protective fishing vessel samples. Whole genome sequencing of the healthcare worker isolates identified the CAL.20C variant of concern (4 isolates 20C/B.1.429, 1 isolate 20C/B.1.427). Patient demographics, symptoms, time to first positive PCR result, Ct value, and time to draw for serology for the healthcare worker cohort are provided in Supplementary Table 6.

## Discussion

Widespread vaccination campaigns are currently underway, and present an opportunity to elucidate correlates of protection from SARS-CoV-2 infection. While many methodologies exist to assess and monitor humoral immunity, the most tractable for clinical laboratory workflows are enzyme immunoassays which can be run at high-throughput with relatively low cost. In this manuscript, we examined the performance characteristics of four EIAs, including the AdviseDx SARS-CoV-2 IgG II assay which was recently granted EUA status, and one surrogate neutralization assay. We found nearly 100% categorical agreement in most comparisons and good correlation between quantitative values, particularly for the anti-S assays. Additionally, we explored how these assays may be used in assessing immune response by examining a cohort of vaccinated healthcare workers who demonstrated vaccine-breakthrough SARS-CoV-2 infection and individuals who demonstrated clinical resistance to infection.

We found the AdviseDx SARS-CoV-2 IgG II assay to have acceptable performance characteristics and strong agreement with four other serologic tests previously granted emergency use authorization. Among a set of 172 patient samples (68 positive, 104 negative), the AdviseDx assay demonstrated a 95.6% sensitivity and 100% specificity. Compared to the manufacturer’s reported sensitivity and specificity of 98.11% and 99.55%, the clinical sensitivity calculated in our study may have been lower due to strict case definitions for infected individuals (14 days post-symptom onset or 10 days following first positive SARS-CoV-2 PCR result) and the inclusion of acutely ill patients (23, 24). Of the three COVID-19 patients who tested negative using this cutoff for the AdviseDx assay, two seroconverted over the following eight days and one died to COVID-19 pneumonia. At concentrations above the manufacturer’s set positive threshold of 50 AU/mL the assay had an acceptable coefficient of variation of less than 5%. While the CV slightly increased at values below the positive threshold, the limit of the blank was calculated to be 17.3 AU/mL, suggesting that negative samples are rarely near the positive threshold and unlikely to cause a false positive result due to analytic variation. In contrast to the Abbott Architect anti-N assay which has demonstrated poor linearity at index values above 3, the AdviseDx demonstrated excellent linearity even when values outside the manufacturer’s analytical measurement range were tested (R^2^=0.998) (25).

The AdviseDx assay demonstrated similar performance to three EUA serological binding assays (Abbott Architect SARS-CoV-2 IgG, EuroImmun Anti-SARS-CoV-2 ELISA, and Roche elecsys anti-SARS-CoV-2 S) measuring SARS-CoV-2 immunoglobulins. For samples collected from patients more than 14 days post symptom onset or 10 days after initial positive PCR result, the AdviseDx demonstrated a 100% positive predictive agreement with all three assays. This high percentage of agreement is in keeping with data from other high-throughput platforms as well (26). A total of 13 discordant samples were identified when categorical agreement across all platforms was assessed regardless of specimen collection date. In two situations, one of the four assays reported a positive value. This occurred twice for the Abbott Architect anti-N assay which reported values near the positive cutoff. Among the other 11 cases, all were AdviseDx positive. Agreement of the other assays with the AdviseDx was as follows: Abbott Architect anti-N (10/11), Roche (8/11), EuroImmun (0/11). Interestingly, the EuroImmun assay, despite having the most instances of discordance, was analytically more sensitive compared to the Abbott Architect anti-N assay when measuring the International Standard (30.2 IU/mL v. 56.2 IU/mL). The discrepancy may reflect a higher relative presence of anti-S antibodies as compared to anti-N immunoglobulins in the international standard. However, other studies have suggested the EuroImmun assay may be more sensitive than the Abbott Architect anti-N (27, 28). It also worth noting that assay sensitivity is heavily dependent on the non-standardized placement of a given manufacturer’s chosen cutoff (29, 30).

The gold standard PRNA method for determining protective antibody titers is time-consuming and requires access to BSL-3 facilities if live SARS-CoV-2 is used (31). While alternative PRNAs using pseudoviruses exist, variation in titer measurements has been demonstrated for different vectors and cell lines (32). As we were unable to compare the results of the AdviseDx SARS-CoV-2 IgG II assay to a gold-standard PRNA or pseudovirus neutralization assay in this study, we instead compared the AdviseDx to a surrogate virus neutralization assay. This assay is currently the only SARS-CoV-2 neutralization assay with EUA approval and has demonstrated excellent correlation with the gold-standard PRNAs (16). Our results demonstrated a positive correlation between anti-S IgG levels and neutralizing values (R=0.86, p<0.001). When regression analysis was performed, we calculated that the 30% neutralizing level (positive cutoff) of the GenScript assay corresponded to value of 107 AU/mL on the AdviseDx assay. This exceeds the manufacturer’s recommended positivity threshold of 50 AU/mL and suggests that when samples are discrepant, they are likely to be AdviseDx positive/ GenScript negative. We observed this trend in our data set with a total of 9 AdviseDx positive/ GenScript negative and 0 AdviseDx negative/ GenScript positive discrepancies. Five of these discrepant GenScript negative results fell between 20%-30% inhibition, and arose from the modification made by GenScript in their positive cutoff from 20% inhibition (in RUO documents) to 30% inhibition in their EUA application (33). The AdviseDx values ranged from 50.1-290.7 AU/mL in discrepant samples. Clinical histories were available in six of the nine discrepant cases. In two instances, samples were collected from patients greater than 14 days after symptom onset. These results suggest the GenScript assay is less clinically sensitive for detection of recent SARS-CoV-2 infection. While the functional data provided by the GenScript assay (i.e. inhibition of ACE2 binding) may better assess for protective antibodies, four of five patients with SARS-CoV-2 infection following vaccination were found to have inhibition levels >89%. These results encourage the cautious interpretation surrogate inhibition assays as markers of sterilizing immunity, especially in the context of emerging variants.

The WHO International Standard for SARS-CoV-2 antibodies was approved in December 2020 and each vial contains 250 IU of the standard (21). We created a dilution series ranging from 100-1 IU/mL and assessed results over five different SARS-CoV-2 platforms. Using the manufacturer’s recommended cutoff values, the Roche was most sensitive at 3.2 IU/mL. The AdviseDx assay had the second lowest threshold for positive detection (8.4 IU/mL) with a 4.6% coefficient of variation. The least sensitive test was the Abbott Architect anti-N (56.2 IU/mL). The IS is prepared at a stock concentration of 1000 IU/mL, however, due to the need to test replicates over a range of instruments, testing was started at 100 IU/mL. This prevented standardization of higher values from assays which were non-linear (i.e. the Abbott Architect anti-N and GenScript).

Patients with innate or acquired immunosuppressive conditions have demonstrated poor antibody responses following vaccination to other viral pathogens (8, 9). Similar trends are being documented following SARS-CoV-2 vaccination (34). Given the severe impact COVID-19 can have on these individuals, longitudinal monitoring of antibody levels may impact decisions related to administering booster vaccinations and altering immunosuppressive regimens. We gathered a small-scale data set of 155 samples from 27 healthcare workers who were vaccinated with mRNA-1273 or BNT162b2 to help characterize what may be considered a typical antibody response. The first positive results were detected on all platforms by three weeks after the first vaccine dose. Antibody levels significantly increased and peaked at 7 weeks, corresponding to the anamnestic booster response. These values may be helpful to physicians as they seek to determine if their patients have demonstrated an appropriate antibody response. Certainly, more work is needed on longitudinal monitoring of antibody levels from vaccination and their functional correlates.

Our study is limited by the relatively small number of patients sampled. While we examined over 100 specimens to determine assay specificity, a larger number should be examined if the AdviseDx assay were to be employed for use in serosurveys. However, test specificity for anti-S serological tests may be less crucial if used to assess serostatus of vaccinated patients. The samples used for determining test sensitivity were primarily obtained from patients who were hospitalized with COVID-19. While seroconversion may begin as early as six days following infection, three patients in our cohort were classified as seronegative when using a cutoff of 14 days post symptom onset or 10 days post first PCR positive in asymptomatic cases. Two of the patients who failed to seroconvert within this timeframe were asymptomatic and eventually seroconverted 13 and 18 days following their first positive SARS-CoV-2 PCR result, indicating anti-S levels are likely associated with the degree of symptoms or infection (24). The third patient who failed to seroconvert died from COVID-19 pneumonia at 17 days post-symptom onset. Several published reports support the view that a cutoff of 14 days post-symptom onset may be too conservative when calculating the sensitivity of SARS-CoV-2 serologic assay. In a series of SARS-CoV-2 infected patients who did not require ICU admission, the 95% CI for seroconversion extended to 25 days post symptom onset (24). Similarly, delayed antibody response at the time of hospital admission for COVID-19 may be associated with poor outcomes (35). These data suggest a longer duration from symptom onset may be appropriate when assessing sensitivity in previously hospitalized or severely ill patients.

During the early phase of the pandemic, serologic assays were used to diagnose patients with prior SARS-CoV-2 infection and to perform seroprevalence studies. However, with the availability of FDA-authorized vaccines and more candidates entering late phase trials, the ability to quantitatively measure the immune response to SARS-CoV-2 may be useful as a biomarker for protection. Such an approach for would be similar to the current practice used in hepatitis B serologic monitoring, where anti-HBs IgG levels above 10 mIU/mL are considered protective (36). To accomplish this task for SARS-CoV-2, studies must be designed to assess the protective threshold. We explored the question of what antibody levels may be considered protective across various platforms through testing samples from three individuals with pre-existing antibodies who demonstrated resistance to multiple SARS-CoV-2 challenges and five individuals who experienced SARS-CoV-2 infection greater than four weeks after receiving their second dose of the BNT162b2 vaccine. Interestingly, we found no difference in the antibody levels between these two groups. It should be noted that the fishing vessel samples were collected prior to the emergence of clinically significant variants. In the vaccine-breakthrough group, all individuals were infected with the California variants 20.C B.1.427/B.1.429. This variant was initially identified in January 2021 and retrospectively found to be in circulation since May 2020 (37). Compared to the wild-type strain, the B.1.427/B.1.429 variants demonstrate an L452R mutation in the RBD and may be associated with increased transmissibility (38). When sera from vaccinated individuals was tested against the B.1.427/B.1.429 variant, it demonstrated a 2 to 3-fold reduction in neutralization compared to the wild type (38, 39). The emergence of novel variants highlights several challenges for diagnostic assays including the difficulty in establishing protective thresholds and impact on the design of capture antigens for EIAs. Though it is exceedingly difficult to profile strain-specific immunity in the context of high-throughput FDA authorized assays given the rapid pace of viral evolution, further work in this space is required to realize more perfect measurements of immune protection.

## Data Availability

data available in summary tables.

## Acknowledgements

The authors thank the staff of the University of Washington Medical Center infection prevention team and virology and immunology clinical laboratories. ALG reports contract testing from Abbott Laboratories and research support from Gilead and Merck, outside of the described work. Abbott had no role in the design or execution of the study.

