## Supplemental Material for "Anti-SARS-CoV-2 antibody levels are concordant across multiple platforms but are not fully predictive of sterilizing immunity"

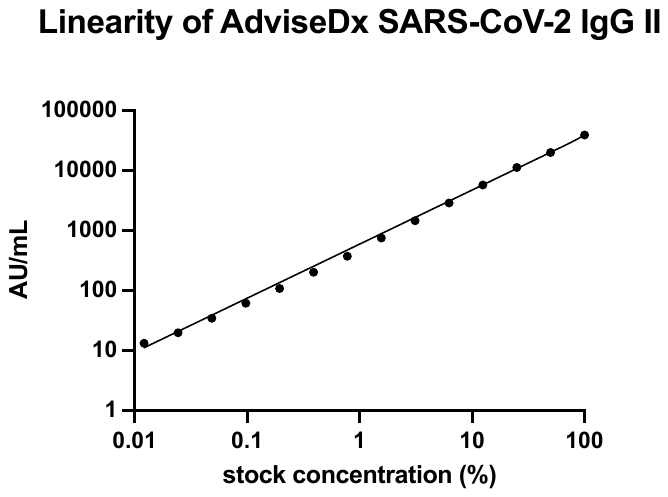

**Supplemental figure 1** The AdviseDx SARS-CoV-2 IgG II assay demonstrates excellent linearity beyond the manufacturer’s recommended analytic measurement interval. A high-positive sample (stock concentration 37,256 AU/mL) was serially diluted two-fold until below the positive cutoff and tested in triplicate. Results demonstrated excellent linearity as measured by the coefficient of determination (R^2^=0.9979).

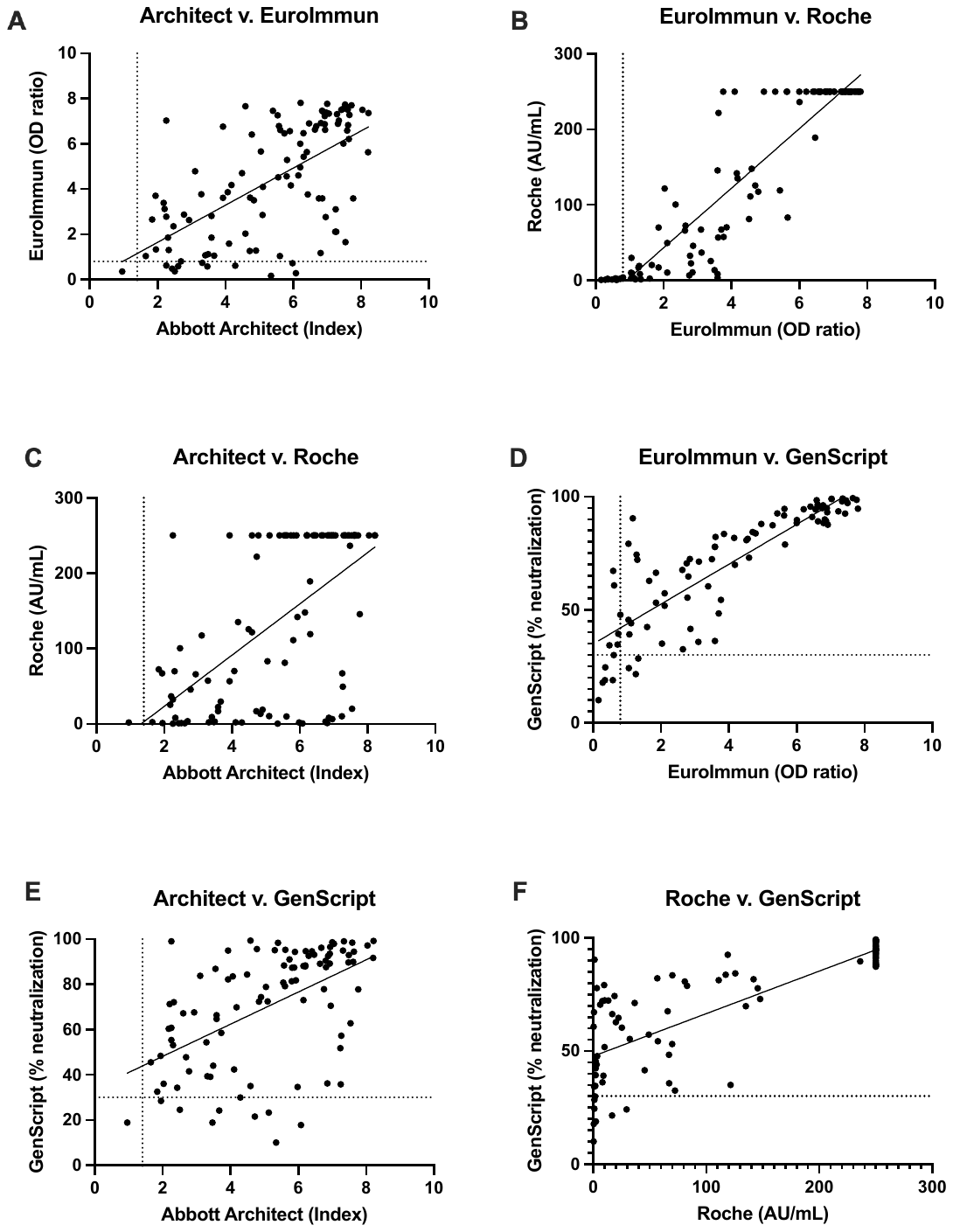

**Supplemental Figure 2** Linear regression plots comparing results from the Abbott Architect anti-N, EuroImmun, Roche, and GenScript surrogate virus neutralizing test assays. The assays demonstrate an overall positive correlation with one another. Goodness of fit (R^2^) for plots A-F are 0.39, 0.85, 0.37, 0.72, 0.28, and 0.62, respectively. Dotted lines represent the positive cutoff value for each assay, cutoff for Roche assay (0.8 AU/mL) not visualized.

|  | SARS-CoV-2  Positive | 2019 Sera (Negative control) |
| --- | --- | --- |
| Anti-S + | 65 | 0 |
| Anti-S - | 3 | 104 |

- 95.6% sensitivity (95% CI: 87.8-98.8%)
- 100% specificity (95% CI: 96.4-100%)

**Supplemental Table 1** The AdviseDx SARS-CoV-2 IgG II assay demonstrates a 95.6% sensitivity and 100% specificity. True SARS-CoV-2 cases were classified as patients greater than 14 days post-symptom onset or greater than 10 days after first positive SARS-CoV-2 PCR result in asymptomatic cases.

|  |  | Dilution | | | |
| --- | --- | --- | --- | --- | --- |
|  |  | Stock | 1:4 | 1:8 | 1:16 |
| Intraday Variation (n=10) | Mean (AU/mL) | 549.2 | 140 | 73.6 | 40.9 |
|  | SD | 17.1 | 4.1 | 3.1 | 2 |
|  | CV (%) | 3.1 | 2.9 | 4.3 | 5 |
| Interday Variation (n=15) | Mean (AU/mL) | 549.7 | 139.5 | 71.8 | 40.6 |
|  | SD | 13.9 | 5 | 2.6 | 3.1 |
|  | CV (%) | 2.5 | 3.6 | 3.6 | 7.7 |

**Supplement Table 2** Intraday and Interday Variation of the AdviseDx SARS-CoV-2 IgG II assay. A four-dilution panel near the positive cutoff was tested ten times by one operator on one day to determine intraday variation. To test interday variation, the panel was tested 5 times per day over three days by two different operators. Under all conditions the assay demonstrated an acceptable coefficient of variation defined as less than 20%.

|  | Abbott Anti-N + | Abbott Anti-N - |
| --- | --- | --- |
| AdviseDx + | 114 | 1 |
| AdviseDx - | 2 | 108 |

- PPA: 98.3% (95% CI: 93.9 to 99.7%)

- NPA: 99.1% (95% CI: 95.0 to 99.9%)

|  | Eurolmmun + | Eurolmmun - |
| --- | --- | --- |
| AdviseDx + | 95 | 11 |
| AdviseDx - | 0 | 52 |

-PPA: 100% (95% CI: 96.1 to 100%)

-NPA: 82.5% (95% CI: 71.4 to 90%)

|  | Roche + | Roche - |
| --- | --- | --- |
| AdviseDx + | 102 | 3 |
| AdviseDx - | 0 | 105 |

-PPA: 100% (95% CI: 96.4 to 100%)

-NPA: 97.2% (95% CI: 92.2 to 99.2%)

**Supplemental Table 3** Categorical agreement between the AdviseDx SARS-CoV-2 IgG II assays and Abbott SARS-CoV-2 IgG (A), EuroImmun (B), or Roche Elecsys Anti-SARS-CoV-2 S (C) enzyme immunoassays regardless of time from symptom onset or first positive PCR result. The positive predictive value of the AdviseDx assay remains near 100%. In the two discrepant cases with anti-N, no other assays were positive, suggesting these results were false positives.

|  | Symptom Onset | Time since PCR pos | AdviseDx | Abbott anti-N | EuroImmun  Anti-S | Roche  Anti-S | Clinical History | Age | Sex |
| --- | --- | --- | --- | --- | --- | --- | --- | --- | --- |
| Patient 1 | 10 days | 2 days | POS | NEG | NEG | POS | Y | 70-79 | F |
| Patient 2 | N/A | N/A | NEG | POS | NEG | NEG | N/A |  |  |
| Patient 3 | N/A | N/A | NEG | POS | NEG | NEG | N/A |  |  |
| Patient 4 | 13 days | 13 days | POS | POS | NEG | POS | Y | 60-69 | M |
| Patient 5 | 21 days | 18 days | POS | POS | NEG | POS | Y | 60-69 | M |
| Patient 6 | 8 days | 7 days | POS | POS | NEG | NEG | Y | 70-79 | F |
| Patient 6 | 9 days | 8 days | POS | POS | NEG | NEG | Y | 70-79 | F |
| Patient 7 | N/A | N/A | POS | POS | NEG | POS | N/A |  |  |
| Patient 8 | N/A | N/A | POS | POS | NEG | NEG | N/A |  |  |
| Patient 9 | N/A | N/A | POS | POS | NEG | POS | N/A |  |  |
| Patient 10 | N/A | N/A | POS | POS | NEG | POS | N/A |  |  |
| Patient 11 | N/A | N/A | POS | POS | NEG | POS | N/A |  |  |
| Patient 12 | N/A | N/A | POS | POS | NEG | POS | N/A |  |  |

**Supplemental Table 4**. Discrepant cases between four EUA enzyme immunoassays for detection of SARS-CoV-2 immunoglobulins. In two situations, low positive Abbott Architect anti-N values and negative results among other assays, suggest these results to be false positives. Among the other discrepant results 11/11 were AdviseDx positive, 10/11 Abbott Architect anti-N positive, 8/11 Roche positive, and 0/11 EuroImmun positive. When available clinical histories demonstrated that discrepant results occurred early (<21 days) during the course of infection. Samples 6 and 7 were obtained from the same patient.

|  | Abbott Architect | EuroImmun | Roche | GenScript |
| --- | --- | --- | --- | --- |
| Abbott Architect |  | 100.0 | 100.0 | 100.0 |
| EuroImmun | 95.7 |  | 95.7 | 96.3 |
| Roche | 100.0 | 100.0 |  | 100.0 |
| Genscript | 91.2 | 96.3 | 93.1 |  |

A.

B.

|  | Abbott Architect | EuroImmun | Roche | GenScript |
| --- | --- | --- | --- | --- |
| Abbott Architect |  | 100.0 | 99.0 | 100.0 |
| EuroImmun | 89.1 |  | 92.3 | 93.9 |
| Roche | 95.4 | 100.0 |  | 98.8 |
| Genscript | 89.2 | 96.3 | 92.0 |  |

**Supplemental Table 5** Positive percent agreement between the Abbott Architect anti-N, EuroImmun, Roche Elecsys Anti-SARS-CoV-2 S, and GenScript surrogate virus neutralizing test assays for patients 14 days post symptom onset or 10 days post first positive PCR result (A) and regardless of time from symptom onset or first positive PCR result (B).

|  | Age (y) | Vaccine | Symptoms | PCR days post-sx onset | CT Value | PCR Platform | GISAID accession | Serology drawn post-symptom onset |
| --- | --- | --- | --- | --- | --- | --- | --- | --- |
| HCW_01 | 60-69 | Pfizer | runny nose, allergy-like symptoms | 3 days | 19.6 | Roche cobas | EPI_ISL_1630160 | 4 days |
| HCW_02 | 40-49 | Pfizer | cough, fatigue, headache, mild congestion | 1 day | 16.4 | Hologic Panther Fusion | EPI_ISL_1490893 | 2 days |
| HCW_03 | 40-49 | Pfizer | cold-like symptoms, loss of taste/smell | 1 day | 17.98 | Abbott Alinity m | EPI_ISL_1497986 | 2 days |
| HCW_04 | 20-29 | Pfizer | stuffy nose, allergy-like symptoms | 1 day | 17.56 | Abbott Alinity m | EPI_ISL_1629715 | 4 days |
| HCW_05 | 30-39 | Pfizer | stuffy nose | same day | 20.8 | Hologic Panther Fusion | EPI_ISL_1601284 | 1 day |

**Supplemental Table 6** Demographic and clinical history for five vaccinated healthcare workers who subsequently developed SARS-CoV-2 infection.
